# Genomic Profiling and Spatial SEIR Modeling of COVID-19 Transmission in Western New York

**DOI:** 10.1101/2023.12.03.23299353

**Authors:** Jonathan E Bard, Na Jiang, Jamaal Emerson, Madeleine Bartz, Natalie A. Lamb, Brandon J. Marzullo, Alyssa Pohlman, Amanda Boccolucci, Norma J. Nowak, Donald A. Yergeau, Andrew T. Crooks, Jennifer A. Surtees

## Abstract

The COVID-19 pandemic has prompted an unprecedented global effort to understand and mitigate the spread of the SARS-CoV-2 virus. In this study, we present a comprehensive analysis of COVID-19 in Western New York, integrating individual patient-level genomic sequencing data with a spatially informed agent-based disease Susceptible-Exposed-Infectious-Removed (SEIR) computational model. The integration of genomic and spatial data enables a multi-faceted exploration of the factors influencing the transmission patterns of COVID-19, including population density, movement dynamics, and genetic variations in the viral genomes replicating in New York State (NYS). Our findings shed light on local dynamics of the pandemic, revealing potential hotspots of transmission. Additionally, the genomic analysis provides insights into the genetic heterogeneity of SARS-CoV-2 within a single lineage at a region-specific level. This interdisciplinary approach, bridging genomics and spatial modeling, contributes to a more holistic understanding of COVID-19 dynamics. The results of this study have implications for future public health strategies, guiding targeted interventions and resource allocation to effectively control the spread of similar viruses in the Western New York region.

## Introduction

The global impact of the COVID-19 pandemic has been profound, necessitating an unprecedented global response to understand, manage, and mitigate the spread of the SARS-CoV-2 virus (1, 2). The novel coronavirus has traversed borders and affected communities on a scale that demands comprehensive research and innovative strategies for public health management(3). Amidst this global challenge, a critical aspect that emerged is the importance of understanding and addressing local transmission dynamics(4). While the broader picture of the pandemic is crucial, the intricacies of how the virus spreads within specific localities is essential for effective public health interventions. Local transmission dynamics not only shape the trajectory of the pandemic but also influence the efficacy of control measures and resource allocation(5, 6). For this analysis, we have chosen to utilize the Western region of New York State, which is characterized by both metropolitan and rural communities with varying population densities. Here we sought to unravel the unique regional factors influencing COVID-19 transmission within these communities.

To develop a comprehensive understanding of COVID-19 dynamics, we employ a dual approach that combines spatially informed SEIR models with detailed genomic analysis of SARS-CoV-2 lineages(7). SEIR models provide a dynamic framework for simulating disease spread based on population movements and epidemiological parameters. In these models, agents occupy various states, including Susceptible, Exposed, Infectious, and Recovered(8, 9). For our model, we first established regional commuter dynamics using state-wide traffic data, followed by more granular census-tract estimations across different social networks (home, work, and school). Our spatially aware modeling strategy allows us to simulate and analyze potential transmission patterns between distinct regions around New York State, accounting for local factors such as population density and commuter movement between neighboring regions(10).

Concurrently, our study incorporates genomic sequencing analysis to investigate the diversity and evolution of SARS-CoV-2 lineages within New York State. By characterizing viral genetic variations at the individual patient level, we gain further insights into the heterogeneity of viral genomes at the sub-lineage level. Thus, the combination of SEIR models and genomic analysis not only enhances our ability to predict and understand the spread of COVID-19 but also provides a unique perspective on how viral genetic variations may contribute to regional differences in transmission dynamics. This integrative approach, bridging computational modeling and molecular epidemiology, offers a robust framework for unraveling the intricate interplay between population-level dynamics and viral evolution in the context of the ongoing pandemic.

## Results

### Statewide variation of major SARS-CoV-2 lineages over time

To first establish whether there were broad regional differences in the SARS-CoV-2 lineages circulating across NYS, we tracked reported caseloads by lineage and location in the Western New York and New York City areas over 2020, 2021, and 2022 (Figure 1). We also included the Canadian providence of Ontario in our analysis due to the close proximity and large commuter population between the southern portion of Ontario (Niagara Falls, Ontario and Fort Erie, Ontario to Buffalo, New York and Niagara Falls, New York)(11). In 2020, we observe distinct lineages between the Province of Ontario and the rest of NYS (Figure 1A). All three regions were high in the B.1 lineage, but by July 2020, deviation in lineages began to take shape, which is likely explained by the strong lock-down measures of the early pandemic. For instance, Ontario saw increased levels of B.1.1 and B.1.1.417, while WNY showed increased levels of B.1.2. and uniquely high levels of B.1.349. Furthermore, during August and September of 2020, Western New York saw increased levels of B.1.349 as compared to Ontario and NYC (Figure 1A). Conversely, lineages B.1.1.434 was specific to New York City highlighting the distinct regional differences at opposite ends of NYS (Figure 1A).

**Figure 1.**
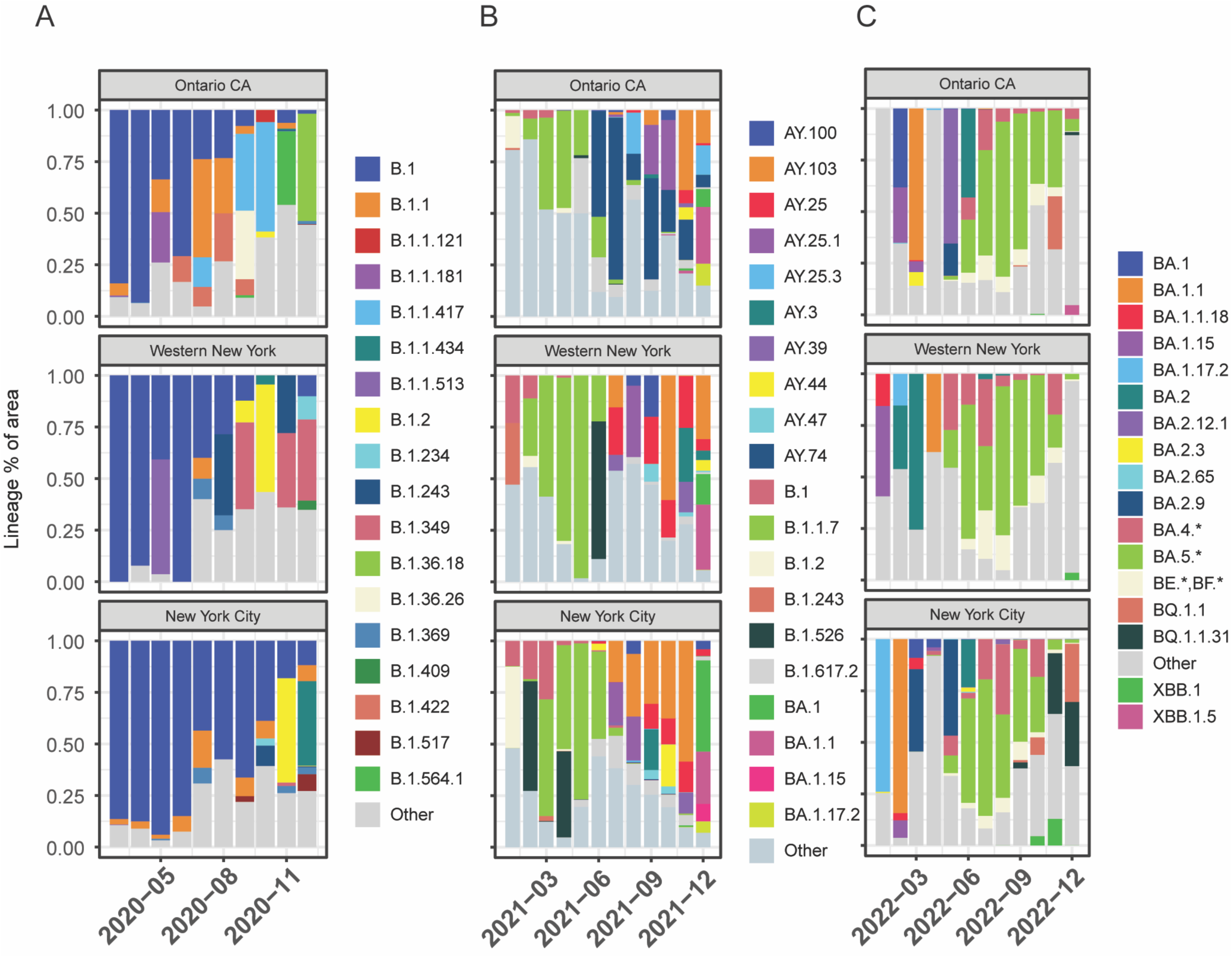
Lineage Distribution of SARS-CoV-2 Across Geographic Regions. Lineage distributions by percentage of total cases within region per month across Ontario, Canada, Western New York, and New York City. (A) 2020 percentages by month (March – December) (B) 2021 percentages by month (January - December) (C) 2022 (January – December)

Next, we were interested in surveying the changing lineage distributions following introduction of major variants of concern to the U.S. Our data suggests that the introduction of Alpha (B.1.1.7) largely replaced the regional differences across NYS, though interestingly did not become dominant in Ontario, and was largely replaced by Delta-based lineages AY.74 by June 2021 (Figure 1B). More heterogeneity is seen following introduction of Delta and its various offshoots (B.1.617.2, AY.103, AY.25 and so forth) (Figure 1C). These results taken together suggest that early in the pandemic, there were distinct, regionally dependent lineages circulating in NYS and these differences were maintained throughout the pandemic, even after introduction of primary variants of concern (Alpha and Delta) at the beginning of 2021. This finding highlights the value of regionally based sequencing efforts for tracking SARS-CoV-2.

Encouraged by our ability to detect regional differences between broad geographic areas, we next sought to analyze the lineage distributions across 10 different economic development regions (EDR) in NYS (Figure 2A). Spanning 2020 – 2022, reported and sequencing viral genomes available in the GISAID database were accessed, and analyzed using rank-correlation coefficient analysis. For each EDR, we correlated the relative rankings to all other EDR, as well as OCA. In 2020, we observed a negative correlation between OCA to all regions in NYS (Figure 2B). Conversely, there were higher correlations between the Mohawk Valley, Central New York, Finger Lakes, and North Country regions, as well as between downstate Mid-Hudson, NYC, and Long Island regions (Figure 2B). In 2021, following decreased lockdown restrictions, the correlation between lineages circulating increased between the majority of NYS EDRs, though OCA was still displaying unique distributions (Figure 2C). Finally, by 2022 we saw a dramatic normalization of the lineages circulating across all EDRs, including OCA (Figure 2D). It is important to note however, that the number of viral genomes sequenced decreased rapidly in the latter half of 2022, resulting in sparse coverage for several EDRs including Mohawk Valley, North Country, and Long Island which may explain slightly correlations to other EDRs.

**Figure 2.**
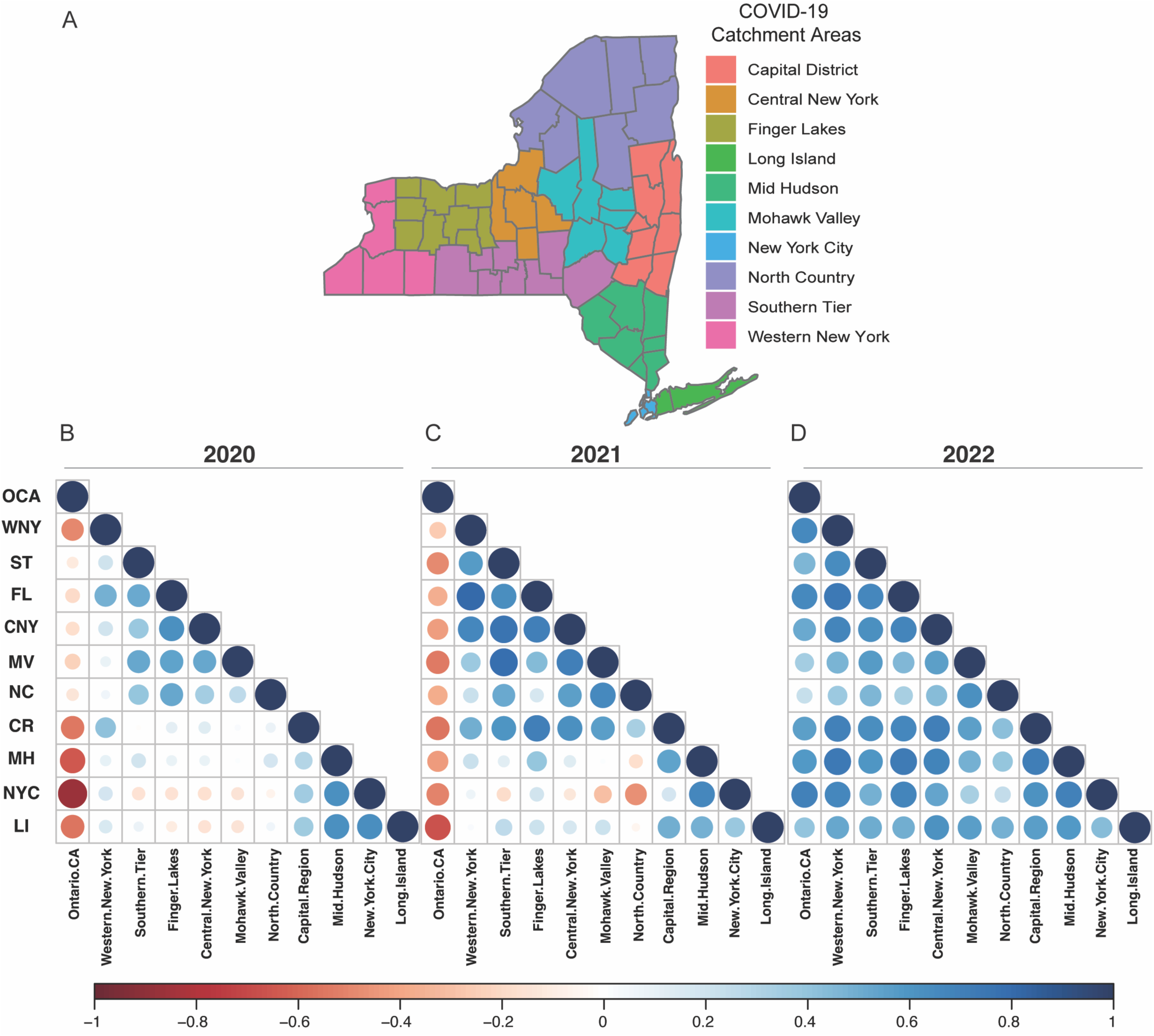
Geographic Economic Development Regions and Lineage Correlations. (A) New York State county grouping into EDR regions. Not shown OCA. (B) EDR Rank-Correlation Coefficient matrix for 2020. (C) EDR Rank-Correlation Coefficient matrix for 2021. (D) EDR Rank-Correlation Coefficient matrix for 2022. All correlations using Pearson correlation coefficient.

### Spatial and Temporal Modelling of SARS-CoV-2 Nucleotide polymorphisms

Based on our findings that there were strong regional relationships on the spread of SARS-CoV-2 lineages across New York State, we next sought to quantify whether we could detect nucleotide level differences in samples belonging to the same variant-of-concern lineage within a single-county in Western New York. As a proof-of-principal analysis, we first evaluated Alpha (B.1.1.7). B.1.1.7 was first introduced in NYS in early December, likely because of air travel between the United Kingdom and one of the New York City Airports(12). Our results indicate that there were additional cases introduced in WNY, however analysis of case numbers per county indicate that B.1.1.7 primarily spread up the Hudson valley and across NYS over a six-month period (Figure 3A). We posited that the B.1.1.7 detected in Erie County then was likely due to multiple introductions, thus there may be distinct nucleotide polymorphisms. To test this, we evaluated 200 B.1.1.7 samples collected in Erie County, New York, between March 2021 and May 2021, and assessed the similarities between viral genomes (Figure 3B). We found that there were several distinct patterns of mutations present in B.1.1.7, lending support to our theory regarding multiple introductions of B.1.1.7 in Erie County (Figure 3C).

**Figure 3.**
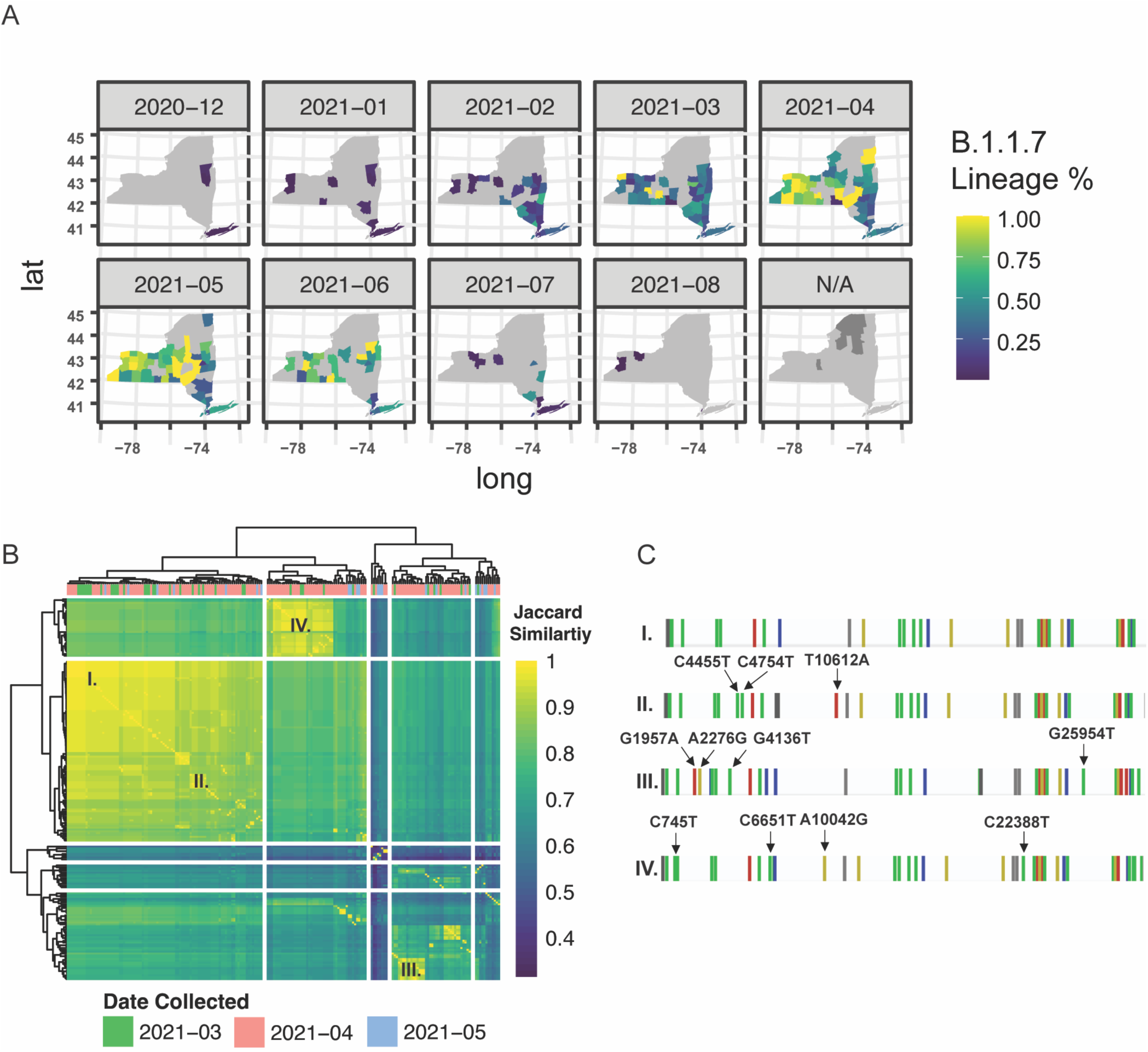
Genetic and Spatial-Temporal Distribution of Alpha B.1.1.7. (A) Geographic introduction and organization of B.1.1.7 lineage from December 2020 to August 2021, by percentage of SARS-CoV-2 circulating in each county per month. N/A represents counties with no B.1.1.7 cases sequenced. (B) Hierarchical clustering of Jaccard distance estimations between variant profiles of 200 Erie Count B.1.1.7 samples. (C) Single base-pair mutational profiles for select B.1.1.7 samples within Erie County.

Encouraged by the analysis of B.1.1.7, we next theorized that there would exist larger mutational differences within a lineage between major metropolitan regions. We next selected Omicron BA.2.12.1 for in depth analysis due to its high virulence and immune evasion potential, as well as robust sample counts across NYS (Figure 4A)(13, 14). Unlike in the case of B.1.1.7, we saw earlier introduction of BA.2.12.1 to Monroe County and Onondaga Counties, where it quickly spread and became dominant in the Central and Western portion of NYS (Figure 4A). Intriguingly, phylogenetic analysis of 2,737 samples from Erie County, Monroe County, Onondaga County, and Westchester County revealed distinct genomic groupings between the different geographically located regions (Figure 3B). Of the samples profiled, Monroe County demonstrates two distinct genomic profiles for BA.2.12.1, further supporting the notion that tracking lineages at the nucleotide level reveals distinct region-specific alterations that are otherwise hidden by broad lineage designations (Figure 3B, C). These results, taken with our analysis of Erie County B.1.1.7 suggests that localized testing of patient samples following by next-generation sequencing analysis provides nuanced distinctions at the genetic level that allows us to examine regional dependencies and relationships between the SARS-CoV-2 lineages circulating at various timepoints.

**Figure 4.**
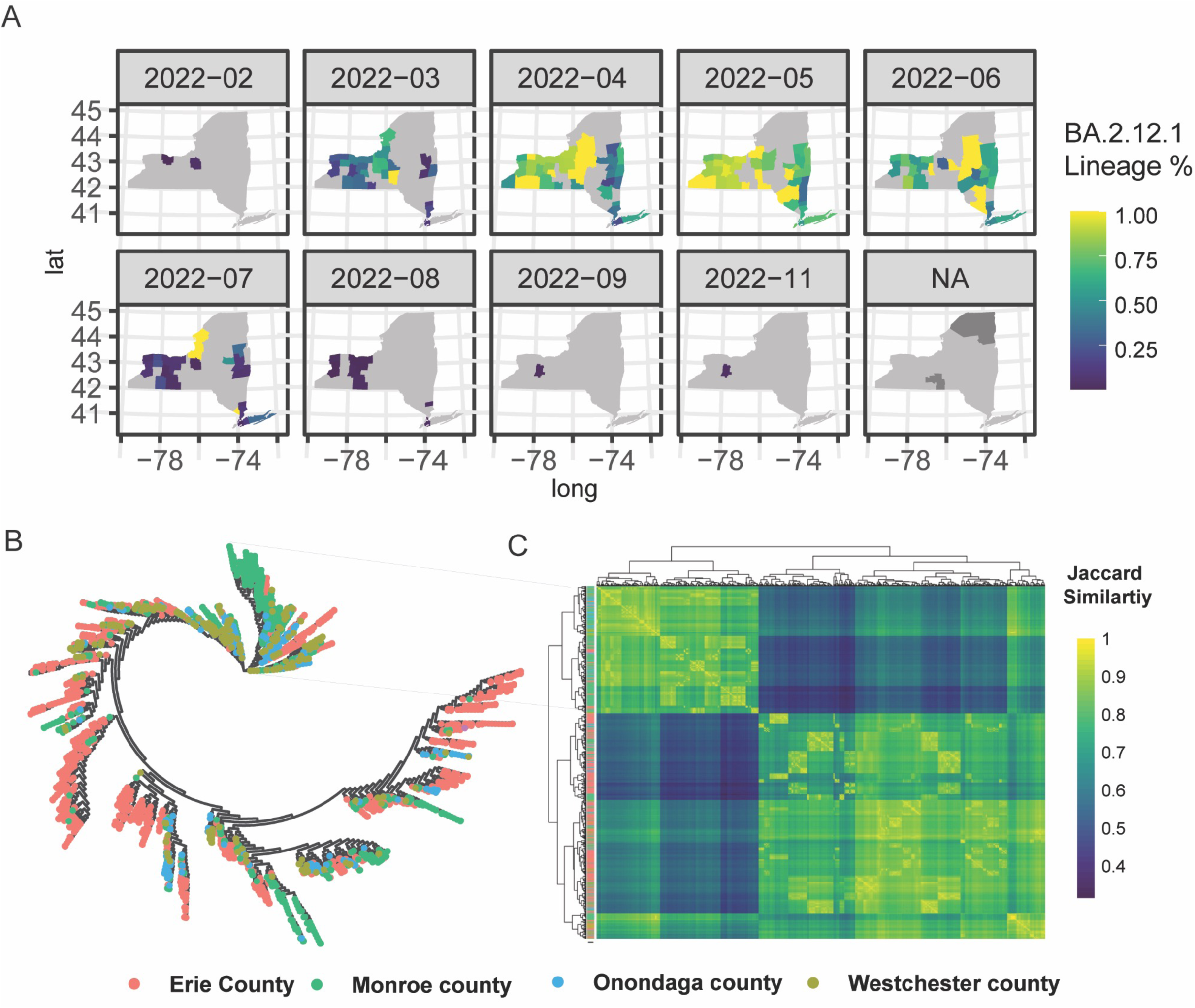
Phylogenetic and Spatial-Temporal Distribution of Omicron BA.2.12.17. (A) Geographic introduction and organization of BA.2.12.1 lineage from February 2022 to November 2022, by percentage of SARS-CoV-2 circulating in each county per month. N/A represents counties with no BA.2.12.1 cases sequenced. (B) Phylogenetic clustering of jukes-cantor distance estimations between consensus sequences of 2,737 samples. (C) Single base-pair mutational profiles for select B.1.1.7 samples within Erie County.

### Establishing Broad Travel Patterns Using Transit Dataset

To better understand the intricate dynamics of COVID-19 transmission in New York State, we sought to examine regional travel patterns, as population movement plays a pivotal role in the spread of infectious diseases. By elucidating the intersections of regional travel and disease transmission, we aim to establish patterns that contribute to region-to-region transmission.

To first establish a general model for large-scale population travel patterns, we leveraged NYS Thruway vehicle traffic data as a proxy to quantify travel behavior which may connect distinct regions across NYS (15). The main-line NYS Thruway spans 426 miles and runs from the WNY region (exit 50) to NYC (exit 15) with extensions into the North Country EDR (Figure 5A). Excluding commercial vehicles, we can detect distinct commuter corridors linking different EDRs. For example, the Capital District Region (Exits 23-26) show increased frequency of travel with the Mid-Hudson (Exits 16-21). (Figure 5B). Alternatively, exits 45 – 47 serve as a hub between the rest of the Western New York region (Exits 48 – 50) as well as increased travel with the rest of the Finger Lakes and Central New York (Figure 5B). Furthermore, specific entrance points, like exit 50 show increased travel that span the length of the thruway, which represent travelers traverse the full extent of the NYS Thruway (Figure 5B). These commuter corridors were consistent with our lineage correlation analysis in 2020 where specific regions showed increased correlations in lineages circulating early in the pandemic (Figure 2B).

**Figure 5.**
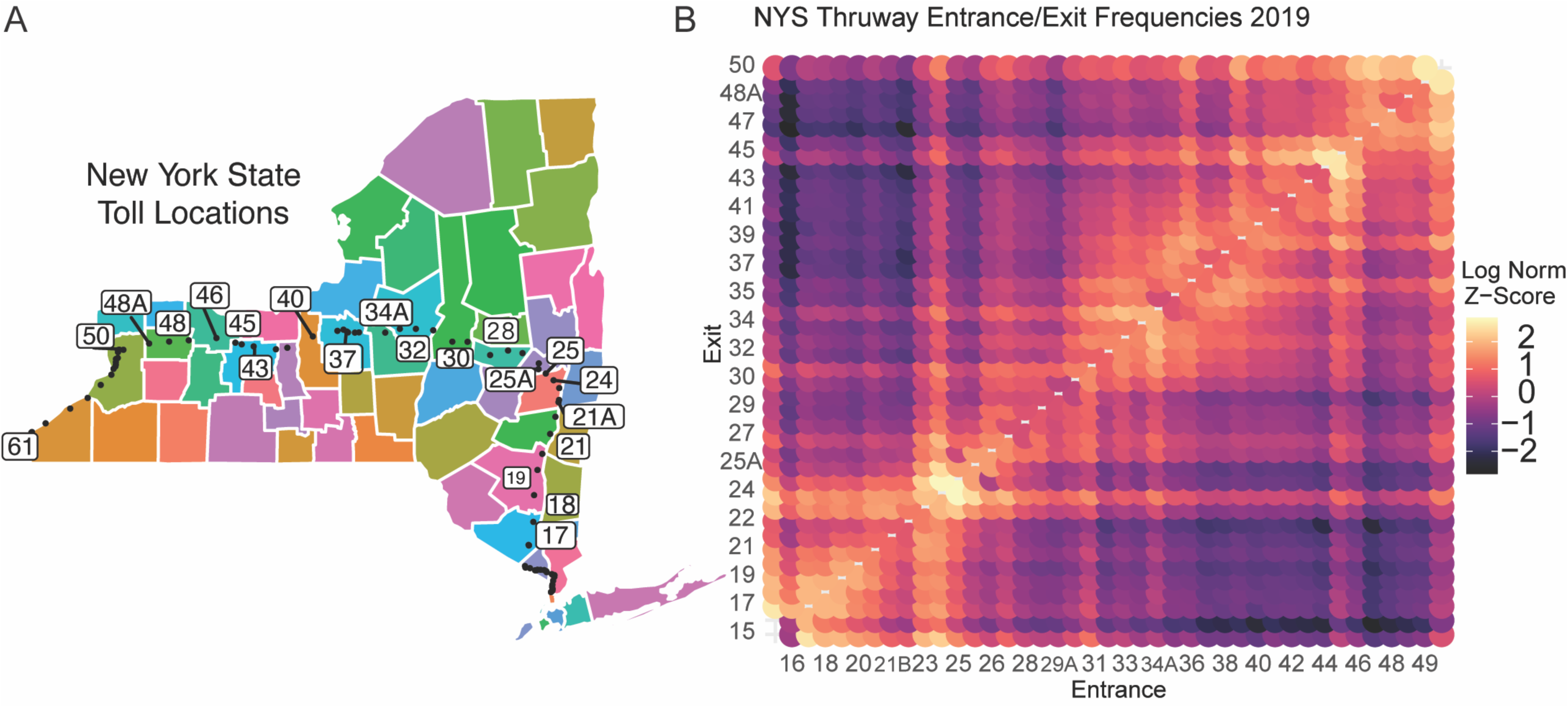
NYS Broad Travel Patterns using Traffic Data. (A)Schematic of NYS Thruway Entrance and Exit points. Black dots represent each toll booth. Numbered exits are subset for visual clarity and regional significance. (B) Log Normalized entrance and exit contact matrix across NYS Thruway. Higher values indicate increased frequency of travel.

### Agent-based Disease Susceptible-Exposed-Infections-Removed (SEIR) Modelling

While the use of the NYS Thruway data was useful to establish broad regional travel dynamics, we next sought to create a more finely tuned model of population-level movement dynamics within Western New York. To accomplish this task, we built an agent-based disease Susceptible-Exposed-Infectious-Removed (SEIR) computational model to simulate how one of the COVID-19 lineages spreads through space and time based on individual (agents) social networks in the Western New York Area (WNY). The purpose of the model is to demonstrate how commuter activity could lead to the diffusion of SARS-CoV-2 in WNY. To build the model 3 steps are involved: first the creation of a synthetic population and its corresponding social networks; second, we build an agent based SEIR model using the synthetic population and lastly, we analyze the simulation results.

As we are dealing with an agent-based model, we first initiated synthetic agents using the approaches previously reported by Crooks et al., 2019(16). To accomplish this initiation phase, we utilized datasets from the US Census for home and work locations and the U.S. Environmental Protection Agency (EPA) for school locations(17). Within the synthetic population, individuals have ages which we break into children (i.e., ages <18) and adults (i.e., ages >= 18). Our model assumes children of school age go to their closest schools or daycares or stay at home with their parents, while adults commute to work or stay at home. The work commute information for adults is consistent with the data from the U.S. Census Bureau’s Longitudinal Employer-Household Dynamics (LEHD) Origin-Destination Employment Statistics (LODES). Full details of the synthetic population generation can be found in previous work by Jiang et, al (7).Then, we constructed social networks (i.e., home, work and education) based on the small-world networks principle(18), where the synthetic individuals are connected based on living in the same household and either working in the same workplace or attending the same daycare/education institute. The rationale for these networks is that an individual might go to work, become exposed to COVID-19, and then go home and infect family members who in turn go to a school and infect students at school, propagating the viral infection through the network. Focusing on Erie County first, we established that two neighboring counties (Niagara and Monroe) saw the bulk of intra-county commuters (Figure 6A). Niagara country residents mainly commuted to Erie County (Figure 6B), while Monroe County served as a major hub for commuters to several different regions, including Erie County, Ontario County, Wayne County, and Niagara County (Figure 6C). The overall inter-connected regional commutes are summarized in Figure 6D. These results suggest that regional transfer of SARS-CoV-2 lineages is likely in the Western New York region due to high levels of daily commuter activity connecting these communities.

**Figure 6.**
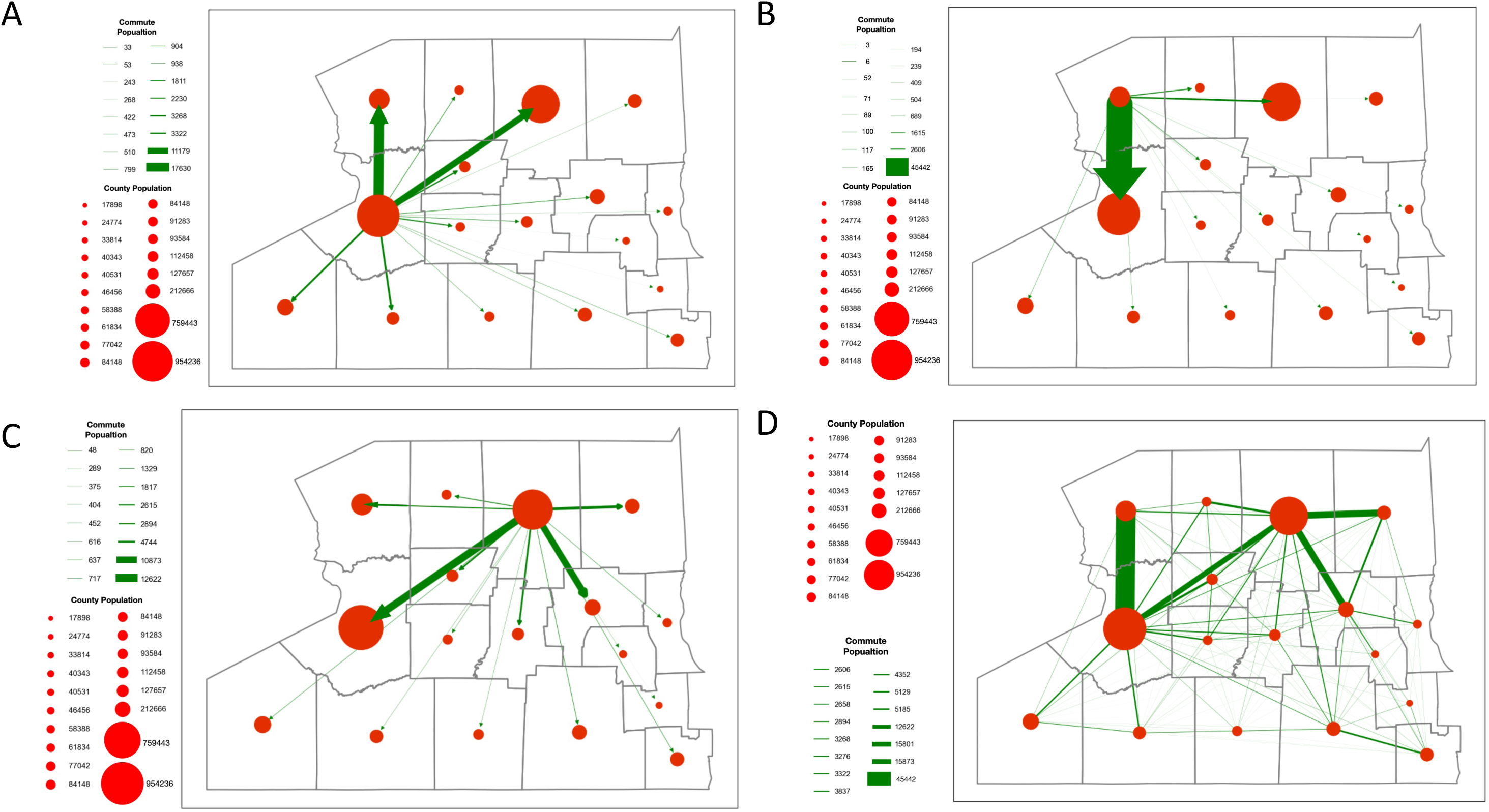
Commuter Behavior Dynamics in WNY. Estimated commuter populations originating in a specific country. (A) Commuter behavior with Erie County origins. (B) Commuter behavior from Niagara County origin. (C) Commuter behavior from Monroe County origin. (D) Composite Commuter behavior network.

As proof-of-principle to see whether our hypothesis that commuter activity could lead to the diffusion of SARS-CoV-2 IN WNY, we next evaluated our (SEIR) model (Figure S1). We simulated the spread of the lineages for 50 days (i.e., 150-time steps) in the Western New York Area. To start the simulation, two agents from Erie County were selected as infected at the start of the simulation. To further analyze the simulation infections result from the agent-based SEIR model, other than reporting the SEIR dynamics, we integrated the results into census tract level and conducted a set of spatial-temporal analyses to demonstrate the diffusion of SARS-CoV-2 lead by commuters in WNY (Figure 7) (19). Within twenty days of introductions within Erie County, our model predicts that several census tracts over 50 kilometers away in Monroe County saw signs of infection (Figure 7B, H). After 30 days and 40 days, there was wide-spread infection across most counties in the Western New York, Southern Tier, and Finger Lakes regions, with a gradient of diffusion around the original infected census tract (Figure 7C-J). Finally, by day 50 our model suggests that all regions in WNY would harbor cases of the lineage introduced into our model, with an increase in cases forming a corridor between Erie County to Monroe County (Figure 7E-I). These results are consistent with our hypothesis that there is strong regional interconnectedness that would fuel the spread from Erie and Monroe county metropolitan regions in a relatively short period of time, and this spread is likely driven by commuter dynamics.

**Figure 7.**
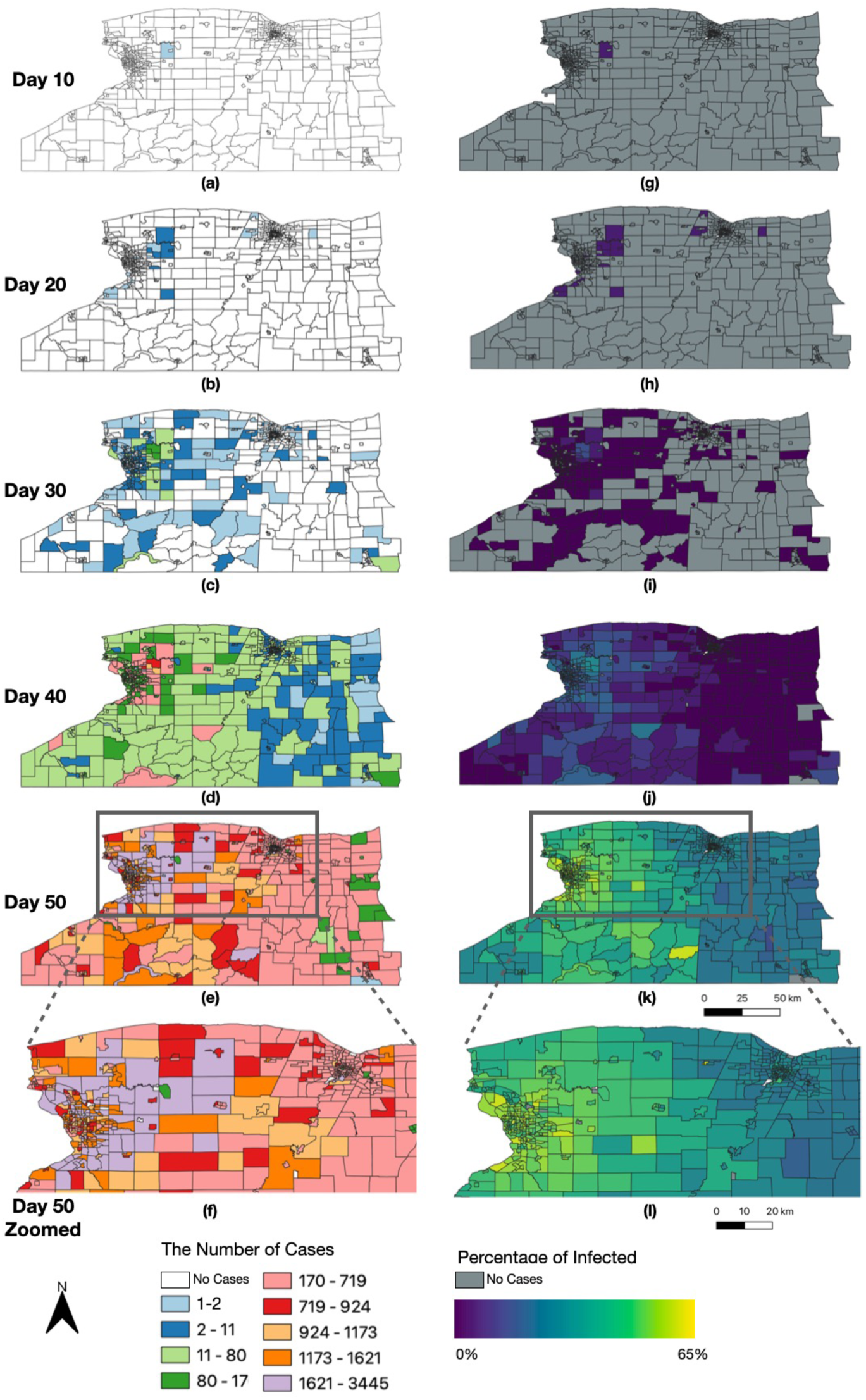
Spatially Resolved SEIR Modelling of WNY. Timelapse of infection case number and by percentage over 50 days, organized by NYS census tract. (A, B, C, D, E, F) Number of total cases since D0 introduction. (G, H, I, J, K, L) Percentage of the census block having been infected with COVID-19 since D0 introduction.

## Discussion

### Regional Monitoring is Key to Understanding SARS-CoV-2 Evolution

In this study, we evaluated regional traffic and commuter patterns and their implications on the genetic diversity and spatial transmission of COVID-19 in New York State, and specifically within Western New York. Our analysis sought to integrate both statewide population movement data, with regionally specific agent-based modelling. We also establish that there are sub-lineage specific mutational patterns in patient sequencing data of SARS-CoV-2. It is our hope that analyses like these will contribute to policy makers’ decisions during future outbreaks and supports the need for continued local and regional monitoring of patient-level viral genomes.

New York State is made up of several dense urban metropolitan centers like New York City, the Capital Region, Utica and Syracuse, Rochester, and Buffalo, with nearly 20 million people as of 2022(20). In between these dense urban centers, are rural communities that our analysis reveals serve as commuting hubs into neighboring counties. Focusing our analysis on the Western New York, our SEIR model suggests the ability for SARS-CoV-2 to diffuse across these rural communities, leading to the transmission from one metropolitan region to another (Rochester, NY to Buffalo NY for instance). While our model did not incorporate vaccination rates, it is also important to note that the counties in between Erie and Monroe tend to have lower percentages of people who have completed the complete course of vaccinations, and also tend to be less likely to have the updated booster formulations(21).

Our genetic analysis of SARS-CoV-2 samples across NYS uncovered regionally specific differences in the genetic backgrounds within specific lineages. While significant resources were invested into regional sequencing hubs, a disproportionate amount of sequencing data was generated downstate in the New York City area as compared to more rural counties, like the Southern Tier and the North Country (Figure S2). Our analysis demonstrates that even within a single SARS-CoV-2 lineage (like B.1.1.7) there exists distinct genetic diversity that could lead to changes in transmission rates at the local level. Due to this, continued investment in infrastructure needed for regional monitoring across NYS should be considered by public policy makers and public health officials.

Despite the prevalence of SARS-CoV-2 infections, sample collection and sequencing of patient-derived samples have decreased since the height of the pandemic. The wide-spread availability of at-home diagnostic tools has reduced collection rates. Furthermore, many pandemic monitoring groups have adopted wastewater-based approaches(22, 23, 24). While wastewater serves as viable tools for measuring overall infection rates, and deconvolution techniques gives an indication of relative proportions of lineages, broad monitoring through wastewater introduces a gap in data for localities without municipal treatment facilities, though recent analysis suggests the feasibility of the method for rural communities (25). Alternative methods, such as SEIR modelling used in this study could serve as potential surrogate strategies.

In conclusion, our study sheds light on the intricate dynamics of the COVID-19 pandemic within the Western region of New York State, emphasizing the importance of understanding local transmission dynamics alongside the broader global perspective. The integration of spatially informed SEIR models and detailed genomic analysis of SARS-CoV-2 lineages provides a comprehensive approach to unraveling the factors influencing transmission patterns. Our analysis of statewide SARS-CoV-2 lineages over time reveals distinct regional differences, especially early in the pandemic. Furthermore, our investigation into single nucleotide polymorphisms within specific VOC lineages at the county level underscores the need for nuanced regional monitoring, exposing localized genomic alterations obscured by broad lineage designations. The establishment of broad travel patterns using transit datasets, and the application of SEIR models demonstrate the interconnectedness of populations and the potential for disease spread across geographic regions. Our findings underscore the significance of regional monitoring, genetic diversity analysis, and spatial modeling in informing public health strategies. As the pandemic evolves, this integrative analysis offers valuable insights for policymakers and health officials to implement targeted interventions, allocate resources efficiently and effectively, and adapt strategies to the evolving landscape of COVID-19.

## Materials and Methods

### SARS-CoV-2 Patient Sequencing Data and Regional Analysis

SARS-CoV-2 viral genomes were accessed and download from them GISAID database for the years 2020-2022, and filtered to New York, United States and Ontario, Canada(26, 27). The date of collection, county, and lineages provided in the GISAID metadata text files were aggregated using the R programming language, and subsequently plotted using the packages ggplot2 (28), lubridate (29), and tidyverse. For the complete GISAID dataset for the spatial analysis of Ontario and New York State, GISAID EPI_SET ID EPI_SET_231204fx <u>10.55876/gis8.231204fx</u>. For the B.1.1.7 analysis, GISAID EPI_SET ID EPI_SET_231204bh <u>10.55876/gis8.231204bh.</u> For the BA.2.12.1 analysis, the GISAID EPI_SET ID is EPI_SET_231204dh <u>10.55876/gis8.231204dh.</u>

### SARS-CoV-2 Economic Development Region Rank-Correlation Coefficients

GISAID metadata from 2020 – 2022 were downloaded and filtered for New York State and Ontario Canada. Location information was post-processed to group by economic development region. The metadata was then collapsed by Date Collection, EDR, and Summation for each Pango Lineage designation. Next, for each EDR we calculated the relative abundance ranking for each lineage and correlated each EDR against all other EDRs. The resulting similarity matrix was next visualized in R.

### Genomic Clustering and Phylogenetic Analysis using the Jaccard metric

Variant profiles for each viral genome were compared to each other using the bedtools jaccard function (30). The jaccard statistic is a similarity coefficient that is defined as the size of the intersection divided by the size of the union of two sets (in this case the variant profiles for each sample being compared). The resulting similarity matrix was used as input into the R pheatmap package for hierarchical clustering and annotated by the county of origin. For phylogenetic analysis, consensus genomes were aligned using the command line version of the MAFFT multiple sequencing alignment algorithm(31). The resulting alignment was then used as input into the FastTree algorithm, inferring maximum-likelihood phylogeny using the jukes-cantor distance model of nucleotide evolution, generating a newick formatted phylogenetic tree (32). For data visualizations, the R packages TreeIO(33), and ggtree(34), using the Pango lineage metadata as data overlays(35).

### NYS Thruway Datasets and Traffic Info

NYS Thruway data was accessed via the data.ny.gov browser(15). Data records for 2019 thruway usage were subset for class 2H and 2L (2-axle vehicles) which corresponds to most non-commercial passenger vehicles. Entries from the database not corresponding to the main-line thruway were also removed. Entrance and Exit points were aggregated over time and the subsequent contact matrix was plotted in R following log normalized z-scoring.

### Spatial SEIR Model

The synthetic population and its social networks are taken as input parameters to initialize the agents of the model. Other parameters related to the lineages (e.g., R0, incubation and recovery period) are also used for the initialization of heterogenous agents, specifically the model assumes a basic reproductive number (i.e., R0) as 3, 7 to 14 days for the incubation period and 4 to 14 for the recovery period(19, 36). Then, SEIR statuses are integrated into the agents to represent their health status. In this model, a time step represents eight hours, where one day is divided into 3 time periods, which are characterized as being home (i.e., either sleeping or getting up), at work (i.e., at work or educational site) and at home (i.e., back at home from work or educational site). The agents interact (i.e., spread the disease or get infected) through their social networks in each time step. In this model, we consider agents who have a work social network as commuters. When these agents are at work, agents will only interact with agents in their work (or school) social network. While agents are at home, they only interact with members from the same household social network, if there is one commuter in the household, the rest of the members of the household have the potential to be infected by the commuter. The model has been programmed to track the overall SEIR dynamics each day (i.e., every 3-time steps). Other than that, the model generates a dataset comprising infectious agents’ information every 10 days (i.e., 30-time steps) during the simulation.

## Supplemental Information

GISAID Identification Numbers for all lineages utilized in this study are in Supplemental Table 1.

## Acknowledgments

This work required collaboration among several entities in the Western New York region, including Erie County Medical Center, Kaleida Health Networks, KSL Diagnostics, Catholic Health, and Erie County Public Health Laboratories. In addition, we graciously thank Wadsworth Laboratories for their initial advisement of our protocol development. The immense work of all laboratories and scientists contributing to the GISAID repository (supplemental tables provided) across New York State proved and invaluable resource for out studies. All work was carried out using University at Buffalo’s Center for Computational Research supercomputing facility, located at the Center of Excellence in Bioinformatics and Life Sciences. This work was initially funded by a State University of New York Research Foundation pilot project award (COVID202044) to J.A.S. Subsequent work was funded by UB’s Genome, Environment and Microbiome Community of Excellence and Erie County Department of Health (J.A.S.) and the National Science Foundation PIPP Phase 1 #2200173 (J.A.S and A.T.C)

## Author Contributions

JEB acquired and analyzed the data, prepared the figures, wrote, and edited the manuscript. NJ acquired the data and developed the SEIR computational model and wrote the manuscript. JE, MB, NAL, BJM assisted with the data analysis. AB, AP and DAY assisted with data generation and provided technical assistance. NJN provided funding and supervised the project. ATC and JAS provided funding, supervised the project, and participated in the manuscript review and editing. All authors contributed to the article and approved the submitted version.

## Data Availability Statement

The agent based SEIR model is shared along with the synthetic population, its social networks at https://figshare.com/projects/SEIR_Western_NY/187872. We do so to allow others to replicate our results and to extend the model as they see fit.

## Figures Legends

**Figure S1.**
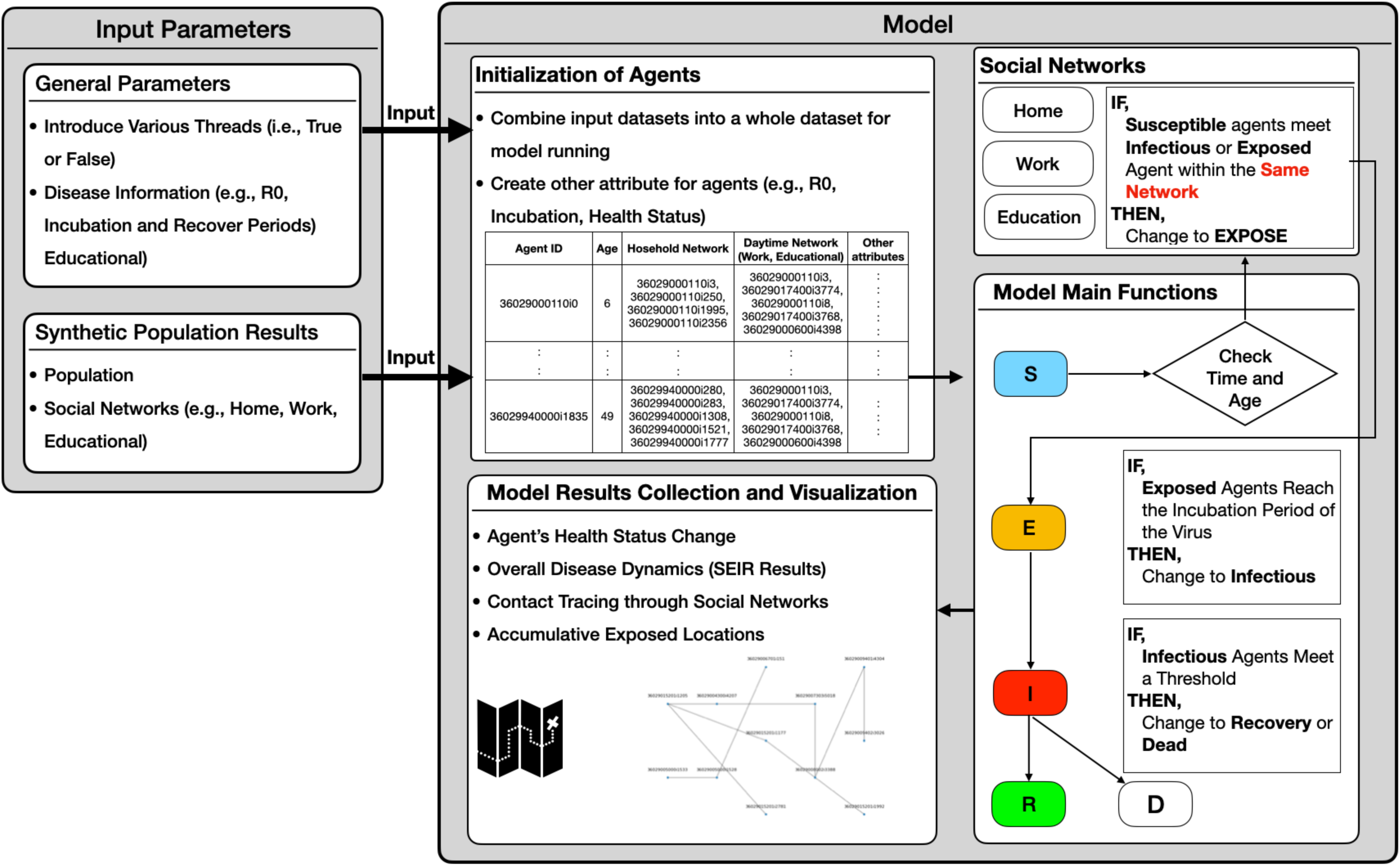
SEIR Model Schematic. Detailed schematic of the SEIR model including general parameter and synthetic population parameter sets, as well as model initialization and function.

**Figure S2.**
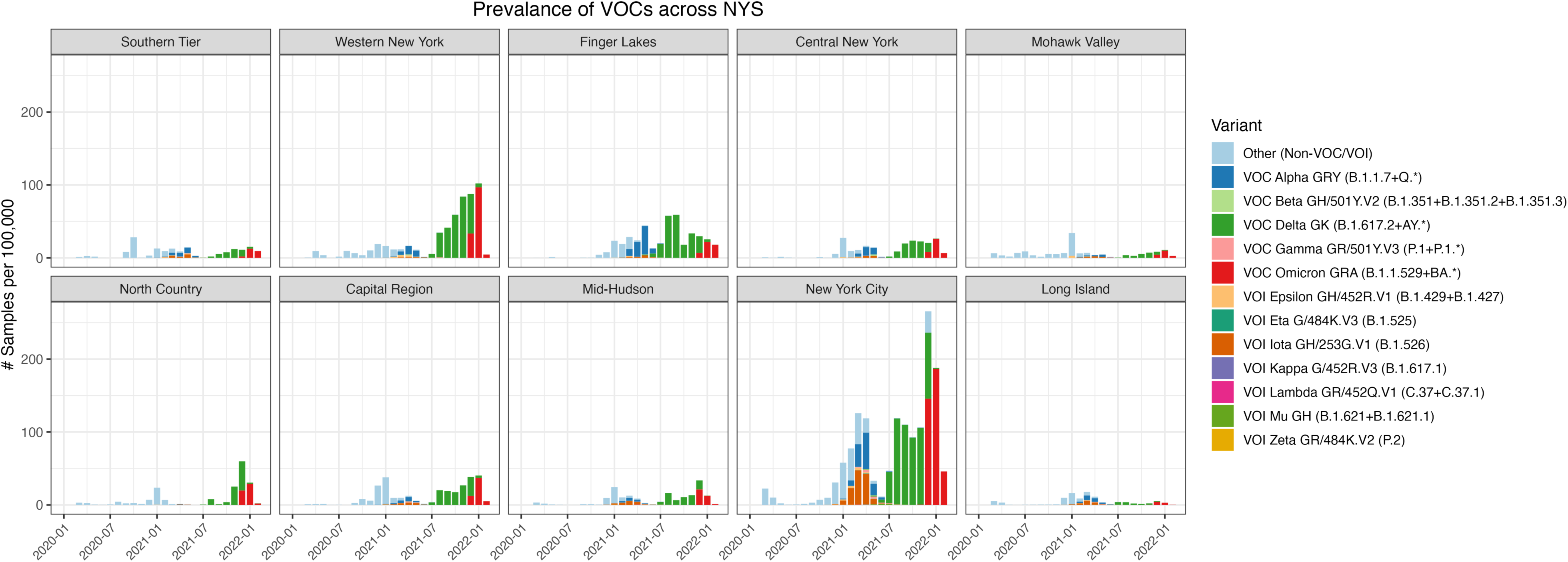
Viral Genomes Sequenced by EDR Reveal Disproportionate Sequencing across NYS. January 2020 to January 2022 total number of viral samples sequenced, organized by EDR, normalized per 100,000 residents.

